# Shotgun metagenomics reveals the microbiome and resistome of water harvesting ponds used by Kenyan rural smallholders

**DOI:** 10.64898/2025.12.05.25341686

**Authors:** B.H. Gregson, A. Bani, L. Steinfield, D. Holt, E.E. Vamos, R. Connell, C. Whitby, R.M.W. Ferguson

**Affiliations:** Applied Ecology Research Group, School of Life Sciences, Anglia Ruskin University, East Road, Cambridge, CB1 1PT, United Kingdom; School of Life Sciences, University of Essex, Wivenhoe Park, Colchester, CO4 3SQ, United Kingdom; School of Science, Aquatic Research Facility, University of Derby, Kedleston Road, DE22 1GB, Derby, United Kingdom; Entrepreneurship & Sustainability, Ivey Business School, University of Western Ontario, 1255 Western Road London, Ontario, Canada; Center for Enterprise and Entrepreneurship, Leeds University Business School, Leeds LS2 9JT, United Kingdom; University of Liverpool (Centre for Genomic Research), Liverpool L69 7ZB, United Kingdom

## Abstract

Water harvesting ponds are essential to smallholder farming across sub-Saharan Africa, yet their role in antimicrobial resistance (AMR) transmission remains unclear. Using shotgun metagenomics, we characterised the microbiome and resistome of 16 rural Kenyan ponds, detecting 582 antibiotic resistance gene (ARG) subtypes across 27 classes. Five ARG types (bacitracin, multidrug, polymyxin, beta-lactam and rifamycin) accounted for most ARGs (90.6%). Genome-resolved analyses recovered 1,542 metagenome-assembled genomes, including non-tuberculous *Mycolicibacterium* carrying rifamycin resistance (*rbpA*) and virulence factors including type VII secretion systems, dormancy regulators, and antigen 85 complex. ARG–mobile genetic element co-localisation was rare, and resistome risk scores were moderate (∼22.4), indicating limited horizontal transfer potential compared to agricultural or hospital effluents. These ponds act as moderate, persistent environmental AMR reservoirs linked to farming practices. Strengthened antimicrobial stewardship, improved manure management and vegetative buffer zones could help mitigate AMR dissemination and support safer rural water systems under a One Health framework.

## 1. Introduction

Water harvesting ponds, fed by rainfall and surface runoff, are an important water source for many rural smallholder farmers across sub-Saharan Africa (SSA), supporting irrigation, livestock and domestic needs in areas lacking piped infrastructure^1,2^. Across the region, tens of thousands of such ponds are in use, with many countries hosting several thousand individually^3^. In Kenya, national and county development programmes have constructed 25,091 household ponds, providing 28 million m^3^ of water storage to irrigate 14,980 acres of farmland^4^. However, their open, untreated nature makes them highly susceptible to contamination from bioaerosols, human activity, livestock and wildlife, particularly during rainfall events when agricultural runoff and faecal material are washed into the ponds^5,6^. Microbiological surveys have reported levels of faecal indicator bacteria and pathogens in harvested rainwater exceeding World Health Organisation (WHO) guidelines, posing risks of diarrhoeal and other waterborne diseases^5,7^. Increasing evidence also suggests that water harvesting environments and agricultural water sources can act as reservoirs for antimicrobial resistance (AMR)^8–10^. Additionally, chemical contaminants such as pesticides may promote AMR through co-selection mechanisms^11,12^. Given the close association between livestock, agriculture and humans, and water resources, these ponds may facilitate the transmission of pathogens and AMR. However, their role as reservoirs of pathogenic and antibiotic-resistant bacteria remains poorly characterised.

In our previous 16S rRNA amplicon sequencing study of 16 water harvesting ponds in Kenya^13^ we found that obligately or facultatively anaerobic bacteria (e.g. *Spirochaeta*, *Opitutus*) dominated the bacterial community. The study also detected the presence of potentially pathogenic bacterial genera, including *Mycobacterium*, *Pseudomonas* and *Neisseria* were detected, and comprised over 1% of the bacterial community, highlighting potential human health risks. While the amplicon sequencing provided a broad overview of community composition, it was limited to genus-level resolution and could not link functional genes, such as antibiotic resistance determinants, to specific microbial taxa^14^. To address these limitations, we extended this work using shotgun metagenomic sequencing to water from the same ponds, enabling strain-level identification^15,16^. Furthermore, this genome-resolved approach allows recovery of metagenome-assembled genomes (MAGs), meaning that taxa carrying antibiotic resistance genes (ARGs), virulence factors and mobile genetic elements (MGEs) can be determined^17^.

## 2. Material and methods

### 2.1. Sample collection

We conducted field surveys at 40 smallholder farms in the County of Laikipia, in Kenya to determine if they had working water harvesting ponds^18^. Of these 40 farms, working ponds were found at 14 sites (Fig. 1). Two farms had two harvesting ponds at the same site (Pond 6/7 and Pond 10/12), meaning 16 ponds in total were sampled. Triplicate surface water (50 ml) was collected from each pond on the same day. All water samples were filtered through a sterile 0.2 μm pore-size syringe filter (Minisar NML, Sartorius). Replicates from each site were filtered using different filters. The volume of water filtered ranged from 6.54 to 15.3 ml. Filters were stored in their plastic casing at −80°C prior to DNA extraction.

**Figure 1.**
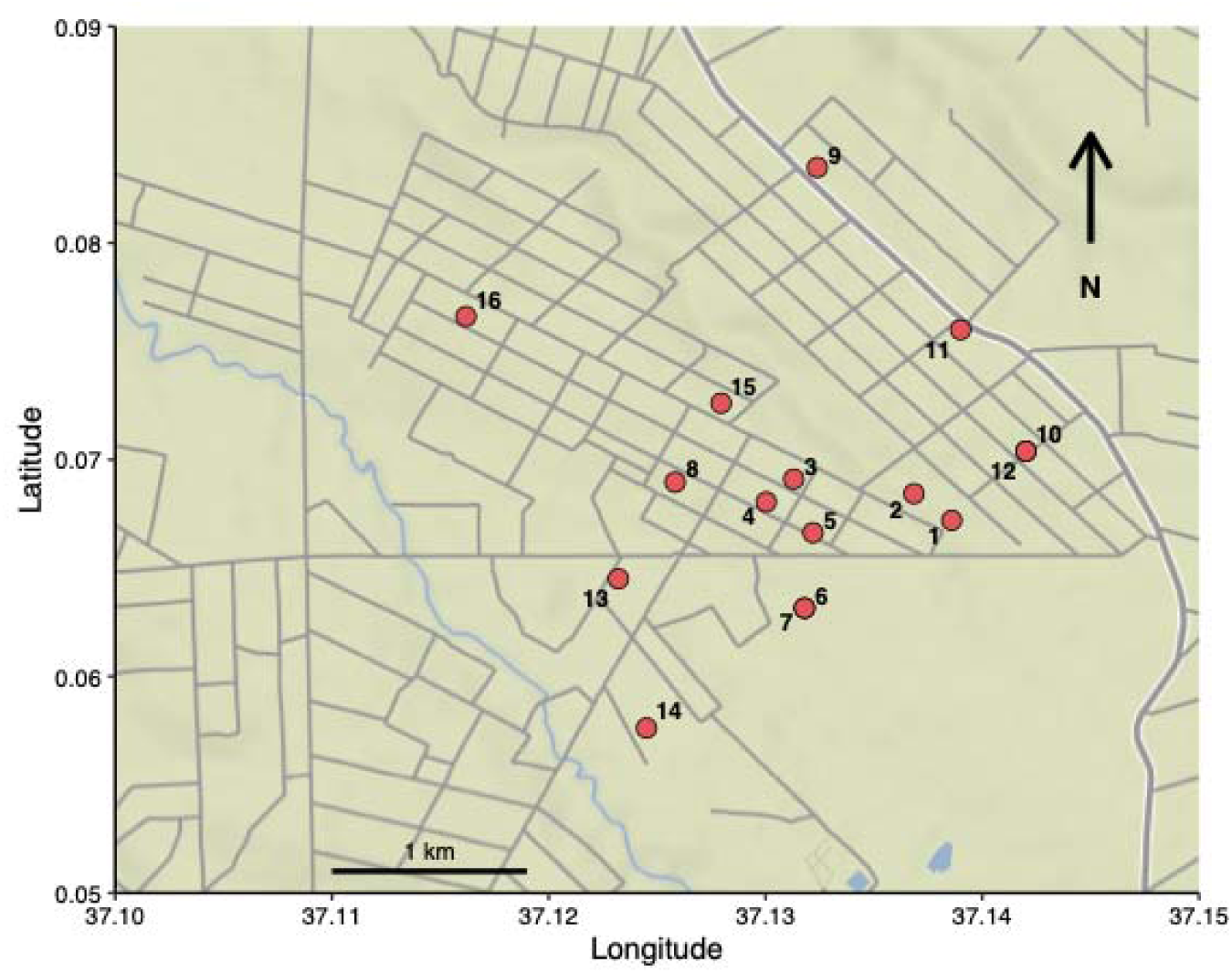
Map showing the location of rural smallholder subsistence farms in the County of Laikipia, Kenya, where surface water was sampled from water harvesting ponds.

### 2.2. DNA extraction

Filters were removed from their plastic casing using a sterile scalpel and tweezers, cut into smaller pieces using sterile scissors and added to PowerBead Tubes (Qiagen) containing 750 μl of PowerBead Solution and 0.7 mm garnet beads. Filters were subsequently homogenised using a Precellys Evolution Homogenizer (Bertin Technologies) fitted with a 2 ml tube holder and white blocking plate (two 30 s cycles at 10,000 rpm). DNA was extracted from homogenised filters using a DNeasy PowerSoil Kit (Qiagen) according to the manufacturer’s instructions and stored at −80°C until use.

### 2.3. Shotgun metagenomic sequencing

Metagenome libraries were prepared using the NEBNext DNA Ultra II FS Kit (New England BioLabs), indexed with custom oligos from Integrated DNA Technologies, and run on an Illumina NovaSeq 6000 using S4 chemistry with paired-end (2 x 150 bp) sequencing at the NERC Environmental Omics Facility (NEOF) at the University of Liverpool. Sequences have been submitted to the NCBI SRA archive under the accession number PRJNA1371399.

### 2.4. Preprocessing of metagenomic reads

The Read_qc module of MetaWRAP ^19^ was used to pre-process raw Illumina sequencing reads in preparation for assembly and alignment. In this module, reads are trimmed with Trim-galore (https://www.bioinformatics.babraham.ac.uk/projects/trim_galore/) and then any human-derived reads (contamination) are removed with bmtagger (ftp://ftp.ncbi.nlm.nih.gov/pub/agarwala/bmtagger/). Read pairs with a single suspected human read are also removed. Reads were normalised to 40x coverage using BBNorm in the BBTools package (https://sourceforge.net/projects/bbmap/).

### 2.5. Identification and quantification of antibiotic-resistance genes (ARGs)

Read-based annotation of ARGs was performed using ARG-OAP v3.0 ^20^. The potential ARG sequences were aligned against the structured antibiotic resistance genes (SARG) database using BLASTX with cut-off values of 10^-7^ E-value, 80% sequence identity and 25 amino acids in alignment length. The ARG abundances were normalised based on estimated cell numbers, shown as ARG copies per cell (copies/cell).

### 2.6. Metagenomic assembly and binning

Co-assembly of the cleaned reads (1,823,007,355 reads) from all samples was performed with MEGAHIT ^21^ generating 42,491,158 contigs. The assembly was formatted to include scaffold length and *k*-mer depth, sorted by length and contigs shorter than 1000bp were removed (6,815,575 contigs remaining). Assembled contigs were binned into three different bin sets within metaWRAP’s ‘binning’ module ^19^ using MetaBAT ^22^, MetaBAT2 ^23^ and CONCOCT ^24^. This clusters together contigs based on sequence properties such as tetranucleotide frequency, differential abundance and similar read coverages across samples. The resulting bins were refined (completeness >50%, contamination <10%) using the ‘bin_refinement’ module within metaWRAP.

### 2.7. Taxonomy and quantification of metagenome assembled genomes (MAGs)

Taxonomy was assigned to the recovered metagenome-assembled genomes (MAGs) using GTDB-Tk (v0.3.2) ^25^. MAG abundances were calculated using Salmon ^26^, as part of the ‘quant_bins’ module in metaWRAP, with default parameters. To further refine the taxonomy assigned by GTDB-Tk, 5S and 16S sequences were extracted from MAGs using Barnnap (https://github.com/tseemann/barrnap). Extracted sequences were compared to those present in the NCBI nr database using the Basic Local Alignment Search Tool (BLAST) ^27^.

### 2.8. Identification of antibiotic resistance genes (ARGs), virulence factors (VFs) and mobile genetic elements (MGEs) on MAGs

Antibiotic resistance genes (ARGs) within MAGs were identified using two complementary tools DeepARG v2.0 ^28^ and the Resistance Gene Identifier (RGI) v5.2.1 from the Comprehensive Antibiotic Resistance Database (CARD) ^29^. DeepARG internally aligns sequences using DIAMOND ^30^ and applies a deep neural network to classify and score potential ARGs based on similarity to known resistance genes. Analyses were performed in gene prediction mode using the long-sequence model, optimised for assembled contigs. In parallel, RGI was applied to protein sequences predicted from each MAG using Prodigal in metagenomic mode ^31^. Proteins were aligned against the CARD database (v.3.3.0) using DIAMOND ^30^ and ARGs were classified based on CARD’s curated homology, SNP and rRNA models. To standardise the outputs from each tool we used hAMRonization ^32^ which converts ARG prediction outputs into a common schema. Predictions from both DeepARG and RGI were filtered to retain only hits with ≥80% sequence identity, generating a unified table of ARG hits including gene identifiers, resistance mechanism, drug classes, alignment scores and associated MAGs.

To identify putative virulence factors (VFs) within each MAG, Prodigal predicted protein sequences were compared against the Virulence Factor Database (VFDB) ^33^ using BLASTp ^34^. BLASTp was run with an e-value cutoff of 1e^-5^ and results were filtered to retain only hit with ≥70% sequence identity. Functional annotations for VFDB protein entries were parsed from the FASTA headers and merged with BLAST results using custom Python scripts. Output files included virulence gene IDs, gene names, functional descriptions, alignment statistics and the corresponding MAGs. To assign virulence categories, a metadata table linking VFDB gene identifiers to virulence categories was joined to the annotated results. A final summary file was compiled containing all virulence factor hits across MAGs, including their functional roles and category assignments.

Mobile genetic elements (MGEs) were identified in each MAG by performing BLASTn searches against curated reference databases for plasmids, phages, transposons, insertion sequences, and integrons ^34^. Custom nucleotide BLAST databases for plasmids and phages were generated using sequences from the NCBI RefSeq plasmid and viral releases, respectively ^35^. Integron sequences were obtained from the INTEGRALL databases ^36^, insertion sequences from ISfinder ^37^ and transposons from the TnCentral database ^38^. Each MAG was queried independently against these databases using BLASTn, with a minimum identity threshold of 90%, a minimum query coverage of 70%, and an e-value cutoff of 1e^-10^.

### 2.9. Resistome risk score

Resistome risk, defined as the potential for ARGs to be mobilised via MGEs and transferred to human pathogens, was assessed using MetaCompare ^18^. Metagenomic reads from each sample were assembled into contigs using the IBDA-UD *de novo* assembler ^39^ and ORFs were predicted with Prodigal ^31^. Assembled contigs with predicted ORFs were annotated against reference databases: CARD for ARGs ^40^; ACLAME for MGEs ^41^; and PATRIC for pathogen associated sequences ^42^. Each contig was then categorised based on whether it contained only ARGs, ARGs with MGEs, or ARGs with both MGEs and pathogen signatures, representing increasing levels of potential public health risk. For each sample, the relative abundance of contigs in each risk category is calculated and used as coordinates in a three-dimensional “hazard space”. A Euclidian distance-based scoring system is then applied to determine the sample’s proximity to the maximum risk vertex (ARG+MGE+Pathogen) and the zero-risk origin, producing a normalized composite resistome risk score, ranging from 0 (low risk) to 100 (high risk).

## 3. Results

### 3.1 Reads-based resistome profiling in water harvesting ponds

Reads-based analysis revealed a total of 582 ARG subtypes, which belong to 27 ARG types, were detected in the water harvesting ponds (**Fig. S1**). The most diverse ARG type was beta-lactam resistance genes, which contained 228 subtypes, followed by multidrug-resistance (88 subtypes), aminoglycoside (54 subtypes), tetracycline (46 subtypes), and macrolide-lincosamide-streptogramin (MLS) (40 subtypes). Five abundant ARG types accounted for 90.6% of the total ARGs (**Fig. 2**): bacitracin (47%, 7.99 × 10^-2^ copies/cell), multidrug (25.2%, 4.29 × 10^-2^ copies/cell), polymyxin (9.7%, 1.65 × 10^-2^ copies/cell), beta-lactam (4.4%, 7.43 × 10^-2^ copies/cell) and rifamycin (4.3%, 7.28 × 10^-2^ copies/cell). *bacA* (bacitracin; 7.91 × 10^-2^ copies/cell), *mexF* (multidrug; 7.57 × 10^-3^ copies/cell), *ugd* (polymyxin; 6.85 × 10^-3^ copies/cell), *rosA* (polymyxin; 5.60 × 10^-3^ copies/cell) and *rbpA* (rifamycin; 5.60 × 10^-3^ copies/cell) were identified as the most dominant ARG subtypes in the water harvesting ponds (**Fig. S2**)

**Figure 2.**
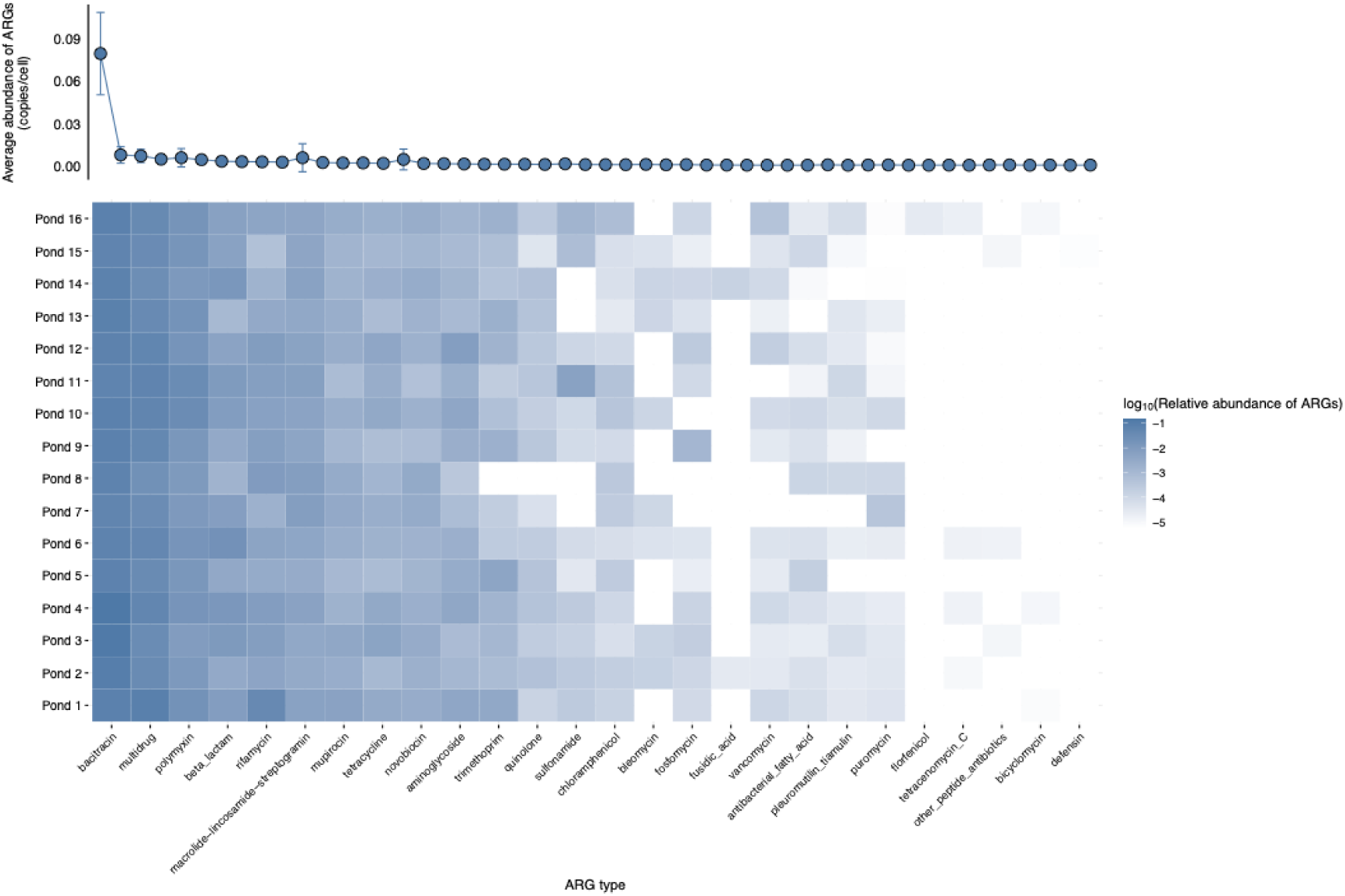
Occurrence and abundance of antibiotic resistance gene (ARG) types detected in surface water samples from 16 water harvesting ponds. Top panel: average abundance (copies per cell) of each ARG type across all samples (±SD). Bottom panel: Heatmap of the log_10_-transformed relative abundance of ARG types across ponds. Darker blue cells indicate a higher relative abundance of an ARG type. White cells indicate ARG types that were not detected. ARG types are ordered by their average abundance across all samples (*n*=48).

We further analysed the occurrence of last-resort ARGs (LARGs) conferring resistance to last-resort antibiotics (carbapenems, polymyxins, vancomycin and tigecycline) in the water harvesting ponds. There were 35 variants of *OXA*, 17 variants of *van*, 1 variant of *KPC*, 13 variants of *mcr* and 39 vairants of *IMP* (**Table S1**). No variants of *tet(X)* were detected in our dataset. Overall, the LARG group with the highest abundance across the ponds was *IMP* with an average of 7.61 copies/cell × 10^-5^ copies/cell, followed by *OXA* (4.03 × 10^-5^ copies/cell) and *mcr* (9.84 × 10^-6^ copies/cell) (**Fig. S3**).

### 3.2. Taxonomic classification of metagenome-assembled genomes (MAGs)

Following co-assembly and binning, a total of 1,542 metagenome-assembled genomes (MAGs) were recovered from the water harvesting ponds, with genome sizes ranging from 0.54 Mb to 13.13 Mb, and GC% contents varying between 26.1% and 74.9%. MAGs had an average completeness of 86.03% (± 8.54%) and contamination of 3.50% (± 2.34%) (**Table S2**). These MAGs were assigned to 32 phyla, 75 classes, 164 orders, 287 families, 586 genera and 82 species (**Fig. S4; Table S2**). The majority of MAGs assigned at the phylum level were affiliated with Pseudomonadota (59.98%), followed by Bacteroidota (14.23%), Actinomycetota (12.09%), Planctomycetota (9.81%) and Verrucomicrobiota (9.16%). The five genera identified with the highest number of classified MAGs were *Asticcacaulis* (2.08%), *Aurantibacillus* (1.17%), *JAFLN01* (1.17%), *ELB16-189* (1.17%) and *M3007* (1.10%). There was a total of 10 classes, 22 orders, 54 families, 108 genera and 331 species that did not have representatives in the GTDB (<95% average nucleotide identity), with the majority of MAGs (1,460) assigned to potentially novel species (**Table S2**).

### 3.3. Antibiotic-resistant bacteria (ARB) and potential pathogens in the water harvesting ponds

Of the 1,542 MAGs, 87 were identified as potentially antibiotic-resistant bacteria (ARB), as they possessed at least one ARG (**Table S3**). These ARGs encoded resistance to 15 classes of antibiotics, with bacitracin resistance being the most abundant (32 MAGs) (**Fig. 3A**). The dominant resistance mechanism utilised by these ARB were antibiotic target alteration (28 MAGs), antibiotic efflux (10 MAGs) and antibiotic target protection (8 MAGs) (**Fig. 3B**). The majority of ARBs were taxonomically assigned to Pseudomonadota (57.47%), followed by Actinomycetota (40.23%). Only a single MAG was taxonomically assigned to Verrucomicrobiota and Bdellovibrinionota. Two MAGs (bin.749 and bin.1325) were predicted to encode the LARG subtype *IMP* and were both taxonomically assigned to the genus *Cellvibrio*.

**Figure 3.**
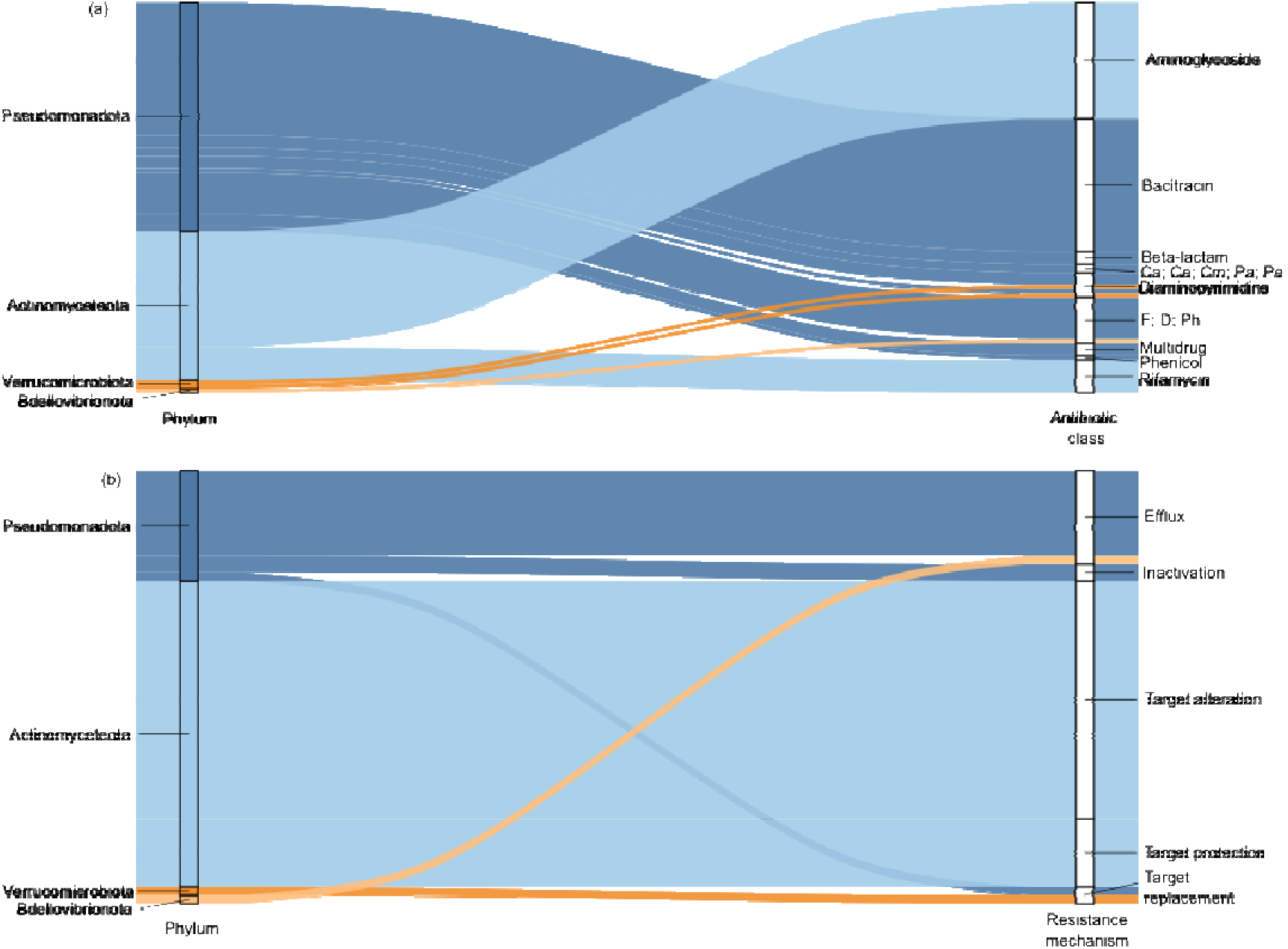
Sankey diagrams showing the distribution of antibiotic resistance gene (ARGs) across bacterial phyla in metagenome-assembled genomes (MAGs) from water harvesting ponds. (a) Flow of ARGs from bacterial phyla to the corresponding antibiotic drug classes. Ca; Ce; Cm; Pa; Pe denotes an ARG that confers resistance to carbapenems, cephalosporins, cephamycins, penams and penems. F; D; Ph denotes an ARG that confers resistance to fluoroquinolones, diaminopyrimidines and phenicols. (b) Flow of ARGs from bacterial phyla to the resistance mechanisms they confer. The width of each flow is proportional the number of ARGs associated with each connection.

A total of 84 MAGs contained both ARGs and virulence factors (VFs) (**Fig. 4; Table S4, S5**). The most abundant MAGs, based on cumulative genome copies per million reads (CPM) across all 16 ponds, were affiliated with the genera *Cellvibrio*, *Mycobacterium, UBA2093*, *Hylemonella* and *Rhodoferax*. *Cellvibrio* (Gammaproteobacteria), a genus of aerobic cellulolytic bacteria commonly found in soil and freshwater, was especially dominant contributing a total of 254.6 CPM across its MAGs. bin.1492 (*Cellvibrio*) was the single most abundant MAG (110.8 CPM) and encoded the highest number of predicted VFs (51). Other *Cellvibrio* MAGs, such as bin.1325 (68.9 CPM) and bin.749 (10.2 CPM), also carried high numbers of VFs (24 and 29 VFs, respectively) and two ARGs each. *Mycobacterium* (Actinobacteria) was the second most abundant genus (124.9 CPM), with bin.1318 (34.1 CPM, 23 VFs) and bin.201 (22.6 CPM, 23 VFs) were among the most abundant representatives.

**Figure 4.**
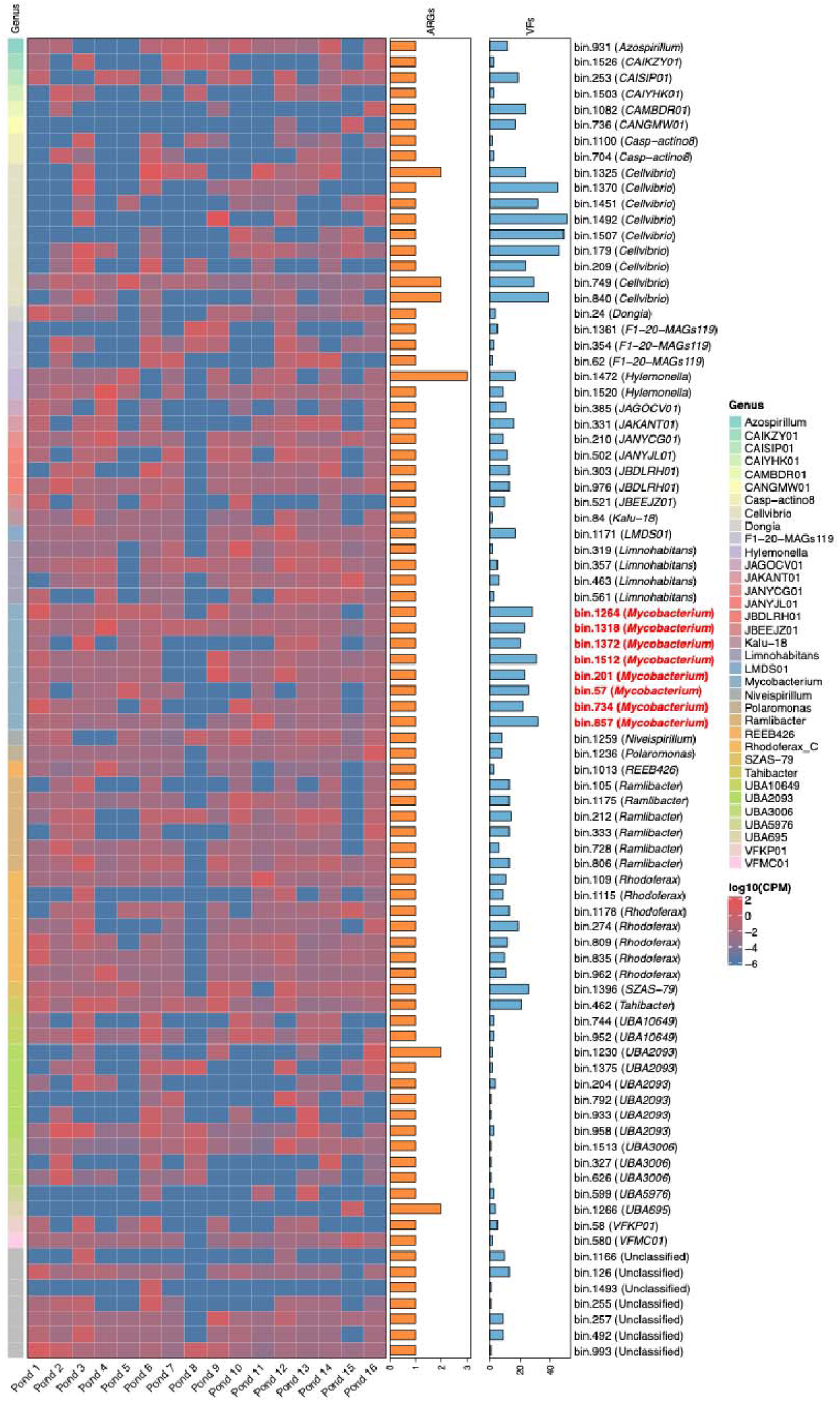
Abundance of metagenome-assembled genomes (MAGs) that contain both virulence factors (VFs) and antibiotic resistance genes (ARGs). MAG abundance is log transformed genome copies per million reads (CPM). Right-side bar plots represent the total number of predicted VFs (orange bars) and ARGs (blue bars) identified in each MAG. Known human pathogenic genera, as defined by ^43^, are highlighted in red.

Other highly represented genera included *UBA2093* (98.7 CPM), a poorly characterised candidate lineage likely representing free-living aquatic heterotrophs; *Hylemonella* (Betaproteobacteria; 60.5 CPM), a genus of motile freshwater bacteria with one known species, *Hylemonella gracilis*; and *Rhodoferax* (Betaproteobacteria; 40.7 CPM), facultative anaerobes within the purple nonsulfur bacteria which are well known for their versatile metabolism including their ability to degrade aromatic compounds. The majority of MAGs (93%) encoded only a single ARG. The highest ARG count was observed in bin.1472 (*Hylemonella*), which carried three ARGs. Virulence factor counts were more variable, ranging from 1 to 51 per MAG and were particularly high in some of the most abundant genera.

Eight MAGs that were originally taxonomically assigned to the genus *Mycobacterium* using GTDB-Tk, based on genome-wide phylogenetic placement. To refine these assignments, 5S and 16S rRNA sequences were extracted using Barnnap, allowing species-level comparisons with reference genomes. All eight MAGs were classified as nontuberculous mycobacteria (NTM) (**Figure 4**). One MAG (bin.57) lacked a detectable marker gene and could not be resolved beyond the genus level. Five MAGs were most closely related to environmental *Mycolicibacterium* species. bin.1318 and bin.1512 shared 98.7% identity with *Mycolicibacterium anyangense* (AP022620.1); a species originally isolated from cattle; bin.1372 had 100% identity to *Mycolicibacterium helvum* (APO22596.1), a species recovered from pitcher plants; and bin.857 showed 98.7% identity to *Mycolicibacterium diernhoferi* (CP080332.1), a soil bacterium. The remaining three MAGs correspond to NTM with known or suspected pathogenicity. bin.201 and bin 1264 both shared 100% sequence identity to *Mycolicibacterium litorale* (CP019882.10), a recently described species implicated in opportunistic human infections^44,45^. Similarly, bin.734 showed 100% identity to *Mycobacterium* sp EPa45 (CP011773.1), a strain whose genome encodes enzymes closely related to those found in pathogenic *Mycobacterium* species^46,47^.

All eight MAGs encoded the antibiotic resistance gene *rbpA* (ARO:3000245), which codes for an RNA-polymerase conferring resistance to rifamycin (**Table S3**). In addition, each MAG contained 20 to 32 putative virulence factor genes (**Table S4**) spanning diverse functional categories including adherence, immune modulation, stress survival, regulation, effector delivery systems and nutritional/metabolic processes. With the exception of bin.1318, all MAGs carried genes encoding the antigen 85 complex (*fbpA, fbpB, fbpC*), a conserved set of mycobacterial adhesins. Multiple regulatory systems known to control mycobacterial pathogenicity were also present. These included two-component systems (*phoPR* in six MAGs; *mprAB* in seven MAGs), the iron uptake regulator *ideR* (six MAGs), and the dormancy survival regulator *dosR* (seven MAGs). Genes involved in host immune evasion were common and included components of the arabinogalactan/lipid biosynthesis pathway, such as *embC* (found in six MAGs). Notably, multiple MAGs carried components of Type VII secretion systems. For example, bin.1512 encoded ESX-1 secretion ATPases (*eccA1*, *eccCa1*/*eccCb1*), bin.857 contained an ESX-3 locus (*eccC3*, *esxG*/*esxH*), and two MAGs (bin.201 and bin.1264) harboured ESX-5 associated genes (*eccb5*/*eccC5* and PE/PPE-family effectors). These NTM genomes harbour a diverse array of virulence factors often orthologous to *M. tuberculosis* virulence determinants, indicating potential pathogenicity within the group.

### 3.4 Mobilome analysis and co-localisation of ARGS and mobile genetic elements (MGEs)

A total of 102 MGEs were identified in MAGs recovered from the water harvesting ponds, including plasmids (*n*=97) and insertion sequences (*n*=5) (**Table S6**). No integrons, phages or transposons were detected. MGEs were detected in 20 MAGs, of which two, bin.857 (*Mycobacterium*) and bin.931 (*Azospirillum*), also harboured ARGs (**Table S3**). bin.857 was the only MAG to contain multiple types of MGE (plasmid and insertion sequences). To assess horizontal gene transfer (HGT) potential, co-localisation analysis of ARGs and MGEs in these two MAGs found no ARGs located within 5kb of MGEs suggesting a low likelihood of ARG mobilisation within these genomes.

To ensure ARGs and MGEs on unbinned contigs were also captured, resistome risk was assessed at the contig level using MetaCompare. All pond samples had MetaCompare resistome scores <25 (mean±SE: 22.42±0.14) placing them in the ‘moderate-risk’ category for aquatic environments as defined by Yang et al. (2025)^48^ (**Table S7**). No significant differences were observed between most ponds, with the exception of Pond 9 (21.35±0.15), which had a significantly lower score compared to Pond 7 (23.61±0.68) and Pond 8 (23.59±0.22) (ANOVA, *F*(15,32) = 3.561, *P* = 0.00125) (**Fig. S5; Table S7**). This elevated risk in Ponds 7 and 8 was associated with a 1.58-fold higher proportion of contigs containing ARGs relative to Pond 9 (**Fig. S6; Table S7**).

To contextualise these results, we compared resistome scores in our samples to values reported for other aquatic environments with established resistome risk rankings (**Fig. 5**). The ponds had an average resistome risk score of 22.4±0.14 which was comparable to values reported for wastewater treatment plant (WWTP) effluent (20.90±0.918). Scores were significantly lower compared to those from agricultural lagoons (25.5±1.00) and hospital sewage (37.8±1.36). These findings indicate that the ponds fall within a moderate resitome risk category, as defined by Oh et al. (2018)^18^ but the absence of ARG-MGE co-localisation within MAGs, and limited detection of MGEs suggest a low potential for HGT in the water harvesting ponds sampled.

**Figure 5.**
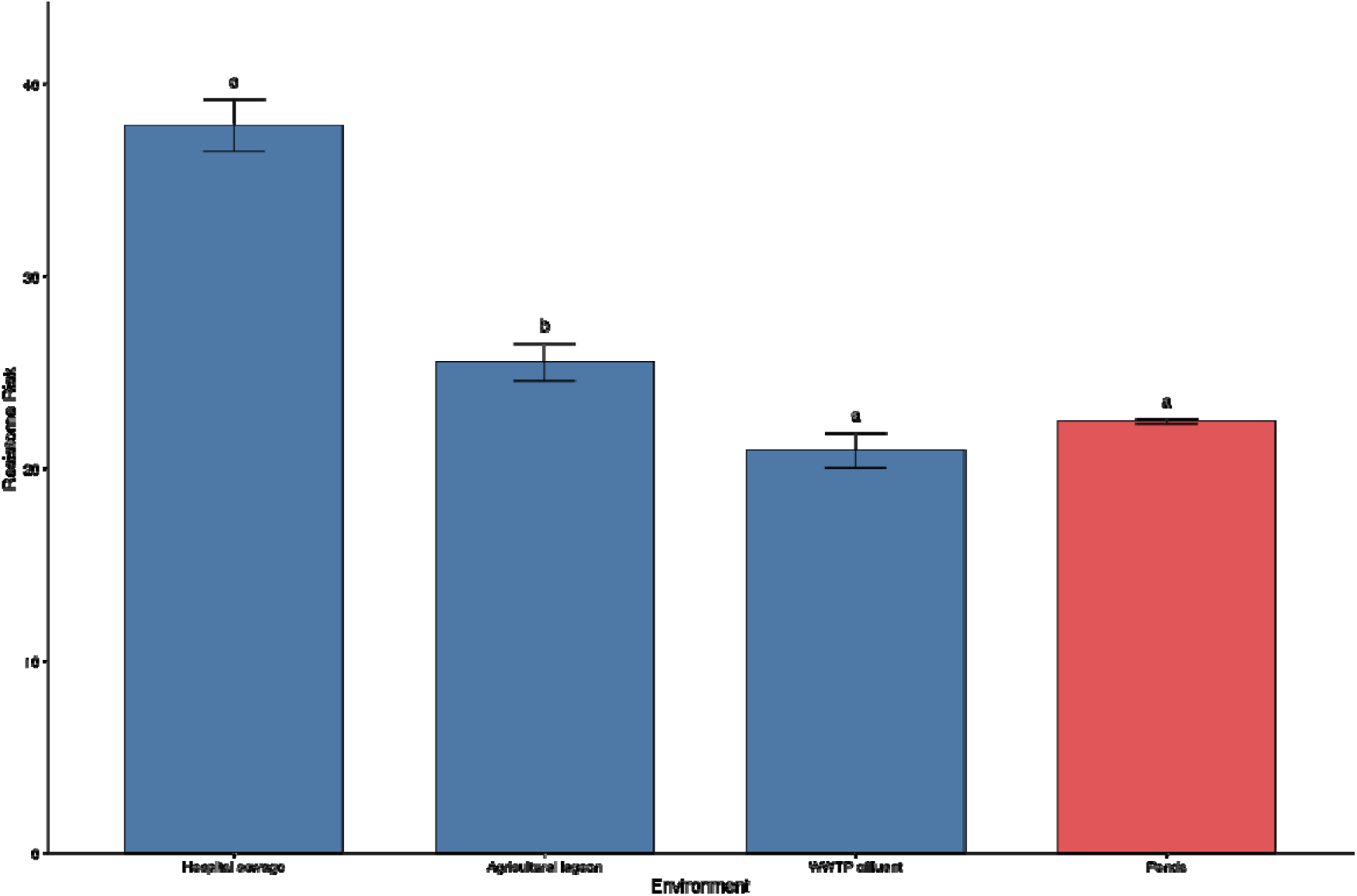
Comparison of resistome risk scores (±SE) in different aquatic with environments with established resistome risk rankings calculated using MetaCompare. The ponds samples in this study are shown in red and compared to other reference environments (blue bars) from Oh et al. (2018)^18^. Bars sharing the same letter are not significantly different based on Tukey’s HSD test (*P* < 0.05).

## 4. Discussion

### 4.1 Antimicrobial resistance (AMR) in water harvesting ponds reflects local agricultural use

Our analysis reveals that antimicrobial resistance patterns in smallholder managed water harvesting ponds reflects the antibiotics commonly used in local livestock farming. Among the 582 ARG subtypes identified, bacitracin resistance genes, particularly *bacA*, dominated the resistome, accounting for nearly half of the total ARG reads across all sampled ponds. This finding aligns with field studies indicating widespread and often unregulated use of bacitracin in Kenyan poultry farming, where it is commonly used as a prophylatic feed additive and growth promoter. Recent nationwide and county surveys found ∼80% of farming households used antimicrobials, with most administering them without vetinary prescription^49,50^. In many rural areas, bacitracin (in the form of zinc bacitracin) is inexpensive, easily accessible through agrovets without a prescription, and considered a routine supplement for broilers and layers. *bacA* is not typically carried on plasmids and is usually a chromosomal, intrinsic gene^51^. Therefore, the detection of *bacA* likely reflects intrinsic bacitracin resistance of the bacterial community, rather than recent acquisition via mobile elements.

Interestingly, resistance to β-lactams, one of the most widely used antibiotic classes in both human and veterinary medicine, was rare in the ponds, comprising <4% of total ARG reads. This is consistent with previous studies that show resistance genes encoding extended-spectrum β-lactamases (ESBLs) are more common in urban areas, but occur at lower levels in rural, low density agricultural environments with reduced pharmaceutical usage^52,53^. For example, in a Kenyan village, only ∼10% of environmental *E. coli* isolates from soil or water samples carried any *bla_CTX-M* variants^54^. The low detection in these rural ponds likely reflects both reduced β-lactam usage in smallholder settings and potential environmental degradation of these compounds before they reach the ponds. Similarly, our study found no evidence of tetracycline destructase genes (e.g. *tet(X)*) or mobile colistin resistance genes (*mcr*), further supporting that antimicrobial inputs into these ponds are relatively restricted to intensively farmed or urban environments.

Across all reconstructed MAGs, only two showed any co-localisation of ARGs with mobile genetic elements, and none showed ARGS integrated within 5 kb of integrases, plasmid markers, or phage sequences. This suggests horizontal gene transfer plays a minor role in these habitats. While this could imply a reduced risk of ARG exchange and spread, it also points to persistent environmental reservoirs that could mobilise under future selection pressures, particularly if antimicrobial use or agricultural runoff increases.

### 4.2 Environmental mycobacteria pose One Health risks

Eight of the MAGs recovered from the water harvesting ponds were assigned to the non-tuberculosis mycobacterial (NTM) genus *Mycolicibacterium*. Like other NTM bacteria, *Mycolicibacterium* spp. are environmentally robust and common across a wide range of environments, for example animal impacted freshwaters^55^ and have also previously been detected in drinking water systems^56–58^. *Mycolicibacterium* spp. are frequently isolated from clinical samples of humans and animals^59–63^, highlighting their significance as an opportunistic pathogen that bridges across all One-Health contexts. *Mycolicibacterium* spp. are commonly resistant to a range of antibiotics, in particular macrolides, beta-lactams, aminoglycosides, and rifampin^64,65^. They have also been shown to have resilience in chlorinated drinking water^57^, making their detection in water harvesting ponds here (that are used for crop watering as well as animal and potential human drinking water) a concern, as removal might be challenging. All the *Mycolicibacterium* MAGs recovered here contained a range of virulence genes (**Table S4**), including Type VII secretion systems (ESX-1, ESX-3, ESX-5) which are implicated in the virulence of *Mycolicibacterium* spp. and previously identified in genomic analysis of *Mycolicibacterium fortuitum*^64^. All MAGs also contained the *rbpA* gene, conferring resistance to rifampin. This gene was also found to be ubiquitous in *Mycolicibacterium fortuitum* genomes investigated from a range of environments (human, soil, fish, companion animal) with samples covering every continent except Europe^64^. Therefore, our study further strengthens evidence that *Mycolicibacterium* spp. have high rates of (or even innate) resistance to rifampin. Across this study MAGs assigned to *Mycolicibacterium* spp. were detected in every pond, with all MAGs containing a range of resistance and virulence genes. Given their ubiquity here, and as outlined previously, their wide-ranging prevalence in the environment (including in drinking water), potential resistance to chlorine treatment, resistance to antibiotics, and records of infections in humans and animals there is a clear risk to health across the One-Health continuum, with water harvesting ponds acting as a conduit for transmission and infection. The increase resistance to rifamycin is also particularly alarming in the context of the usage of these antibiotics to treat severe cases of carbapenem-resistant bacterial infections^66^. In the last few years, Kenyan hospitals have seen an increase of carbapenem-resistant infection between 7-17% with a third of the cases classified as difficult to treat^67^. It should be noted that a further challenge to mitigating this risk is that *Mycolicibacterium* were recently reclassified from the *Mycobacterium* genus (e.g. *Mycobacterium tuberculosis*)^68^. The reclassification of *Mycobacterium* into five new genera may be justified based on genomic analysis but has the potential confusion^69^ in clinical and regulatory contexts.

### 4.3 Policy and on-farm mitigation strategies

At the policy level, strengthening Kenya’s existing antimicrobial stewardship framework is critical. The National Action Plan for AMR outlines strategic objectives, including improved governance, strengthened multisectoral coordination, enhanced surveillance, and optimisation of antimicrobial use through education, stewardship and regulation^70^. Whilst this plan prohibits the use of antibiotics for growth promotion and mandates prescription-only access to veterinary drugs, implementation remains inconsistent, particularly in rural settings where over-the-counter sales and informal distribution persist^49^. Enhanced enforcement through licensing and inspection of agrovet outlets, and improved monitoring of antibiotic imports, could help curb the unregulated distribution of bacitracin and other antibiotics commonly used by smallholders. Expansion of integrated One Health AMR surveillance systems, encompassing livestock, humans and environmental compartments, such as water harvesting ponds, would provide early warning of resistance gene dissemination^71,72^. The National Action Plan also emphasises awareness creation, risk communication and behaviour change as key targets for mitigating AMR, recognising that farmer education and capacity building is key. Targeted training through county agricultural extension officers, community health workers and County AMR Stewardship Interagency Committees could improve prudent antibiotic use and waste management practices^49,70^.

At the farm scale, practical interventions can directly reduce ARG transmission into the water harvesting ponds. Controlled aerobic composting of livestock manure can significantly reduce the abundance and diversity of ARGs before land application^73,74^. Aerobic composting not only reduces viable antibiotic-resistant bacteria but can also degrade residual antibiotic compounds^75,76^. Although an unintended consequence of this could be increased exposure to bioaerosols and associated health effects, as composting is known to emit vast quantities of health relevant bioaerosols, including *A*s*pergillus fumigatius,* and many bacteria implicated in AMR^77^. Aerobic composting is technically feasible for rural subsistence farmers when adapted to low-cost, small-scale systems. Simple compost heaps or passively aerated windrows have been recommended as a low-cost option for African countries^78^. Farmers can layer livestock manure with dry plant biomass to balance C:N ratios and turn the pile at regular intervals with simple tool (e.g. fork or hoe) to maintain aeration^79^. Maintaining aerobic conditions have been shown to substantially reduce ARG loads during composting^80–82^. Promoting low-cost composting facilities or cooperative manure treatment systems among smallholders would therefore mitigate ARG inputs into surrounding water bodies.

Additionally, vegetated buffer strips could be installed between the ponds and adjacent agricultural land to intercept surface runoff containing microbial contaminants, manure particles and residual antibiotics. Vegetated buffer strips are bands of permanent, dense plant growth positioned to intercept runoff from an upland area, such as an agricultural field^83,84^. These strips have been shown to be effective at reducing the transport of manure associated resistance contaminants off agricultural fields^85,86^. Similar approaches could be adapted for smallholder farms in Kenya, where narrow grassed strips or locally adapted vegetation (e.g. Napier grass, Vetiver grass) can be planted along pond perimeters or drainage paths.

The design of the water ponds promoted by the NGOs that support their construction (through funding and training) for small-holder farms recommends a screening around the ponds of bushes or branches as a safety measure, preventing animals and children falling into the ponds. Only one of the surveyed farms had such a barrier and attention should be paid to promoting the uptake of these vegetation barriers around the ponds. Fencing and providing alternative watering points can prevent the transmission of ARBs and ARGs from livestock into harvested water systems. Installing livestock exclusion fencing around ponds has been shown to reduce faecal deposition, sediment disturbance and bacterial contamination of surface waters^87,88^. Providing alternative watering points, including troughs or storage tanks, positioned at a distance from the pond, would help divert livestock away from pond edges and reduce the likelihood of direct contamination through trampling or defecation^89,90^.

Together these policy and farm-level measures offer a strategy for mitigating AMR dissemination in rural Kenya agroecosystems. Strengthening enforcement, education and surveillance at the national level, combined with composting, vegetative buffers and livestock management at the farm level, can collectively reduce the environmental loading of resistance determinants, such as *bacA* and *rbpA*, supporting sustainable water and food security under the One Health framework.

### 4. 4. Conclusion

In conclusion, this study provides one of the first high-resolution resistome profiles of smallholder managed water harvesting ponds in sub-Saharan Africa. We show that while overall resistome risk is moderate, these ponds consistently harbour ARGs linked to agricultural use, particularly *bacA*, conferring resistance to bacitracin. The identification of multidrug-resistance and virulence-associated *Mycolicibacterium* spp. underscores the potential for these water systems to act as environmental reservoirs and transmission points for opportunistic pathogens. Although mobile genetic elements and B-lactamases were rare, the persistence of ARGs in core environmental taxa highlights the role of unmanaged agricultural runoff and livestock in shaping aquatic microbiomes.

As water harvesting ponds continue to expand across drought prone regions of Africa, minimising the presence of pathogens and AMR is crucial. Practical low-cost interventions, such as composting livestock waste, installing buffer vegetation and better management of livestock should reduce the presence of ARB. At the policy level, sustained investment in antimicrobial stewardship, environmental surveillance and farmer education will be essential to reduce AMR. Our findings demonstrate the value of integrating environmental metagenomics with One Health approaches to inform policy, improve rural water management and reduce AMR transmission risks in smallholder farming systems.

## Funding

This research was funded by the NERC Environmental Omics Facility (NEOF) Pilot Project Competition (NEOF1385) and the Global Challenges Research Fund (GCRF) ‘Investigating the effectiveness of a low technology planter for subsistence farming’ (Project A004, GCRF74). The NERC Environmental Omics Facility (NEOF) is funded by the UK Natural Environmental Research Council (NERC).

## Data Availability

All data produced in the present study are available upon reasonable request to the authors

## Notes

### Competing Interest Statement

The authors have declared no competing interest.

