## Supplementary figures and images for "Shotgun metagenomics reveals the microbiome and resistome of water harvesting ponds used by Kenyan rural smallholders"

### Supplementary Figure 1

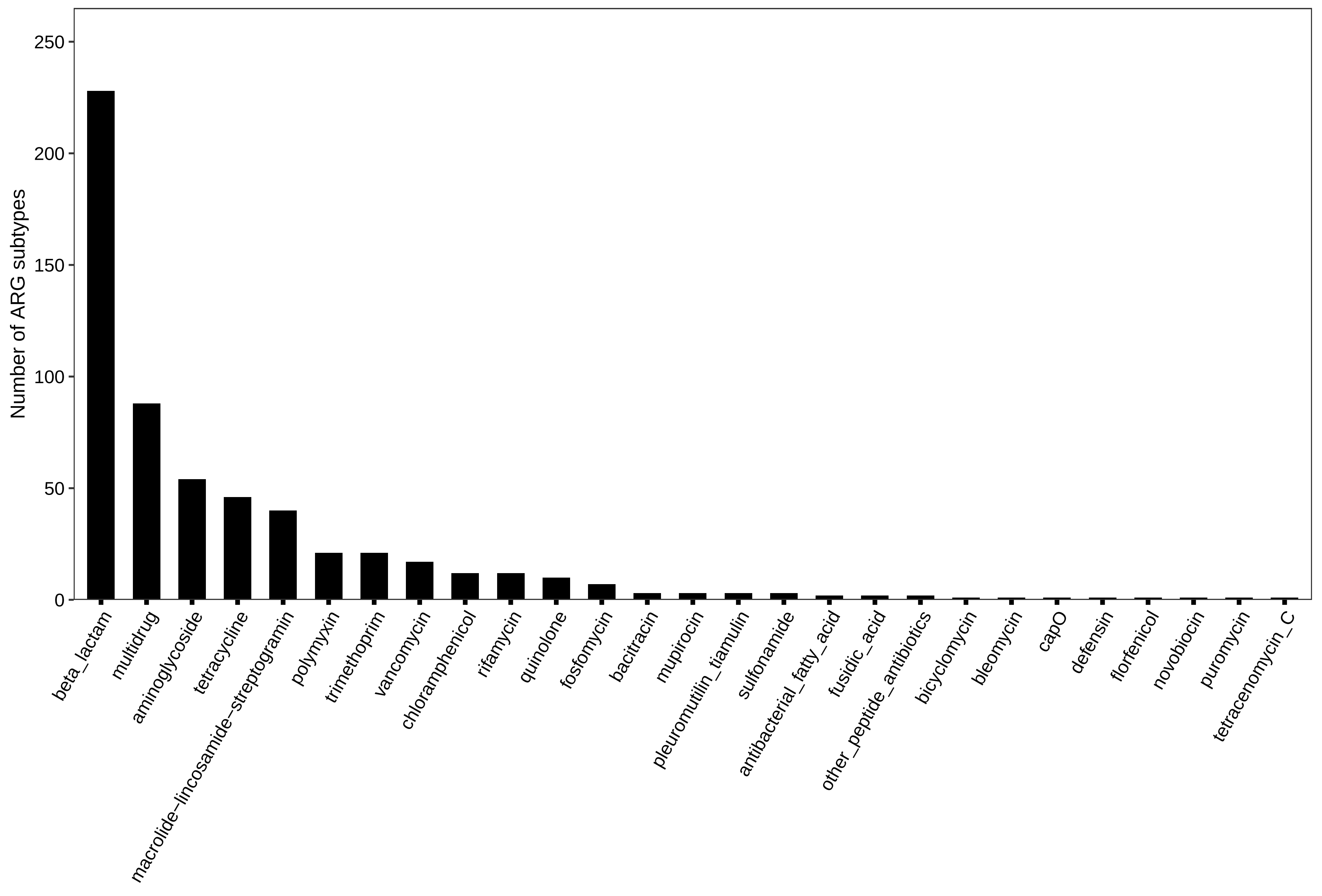

### Supplementary Figure 2

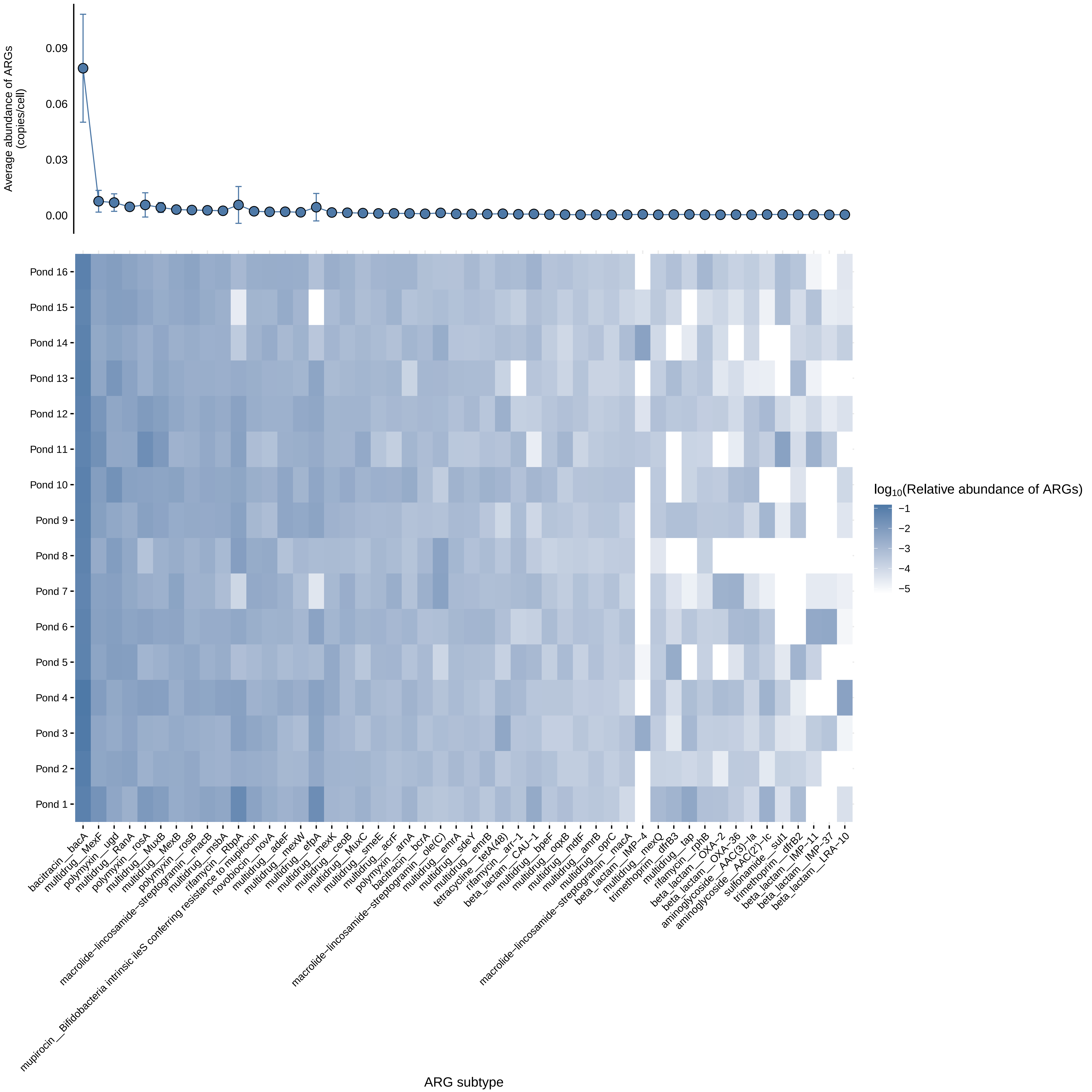

### Supplementary Figure 3

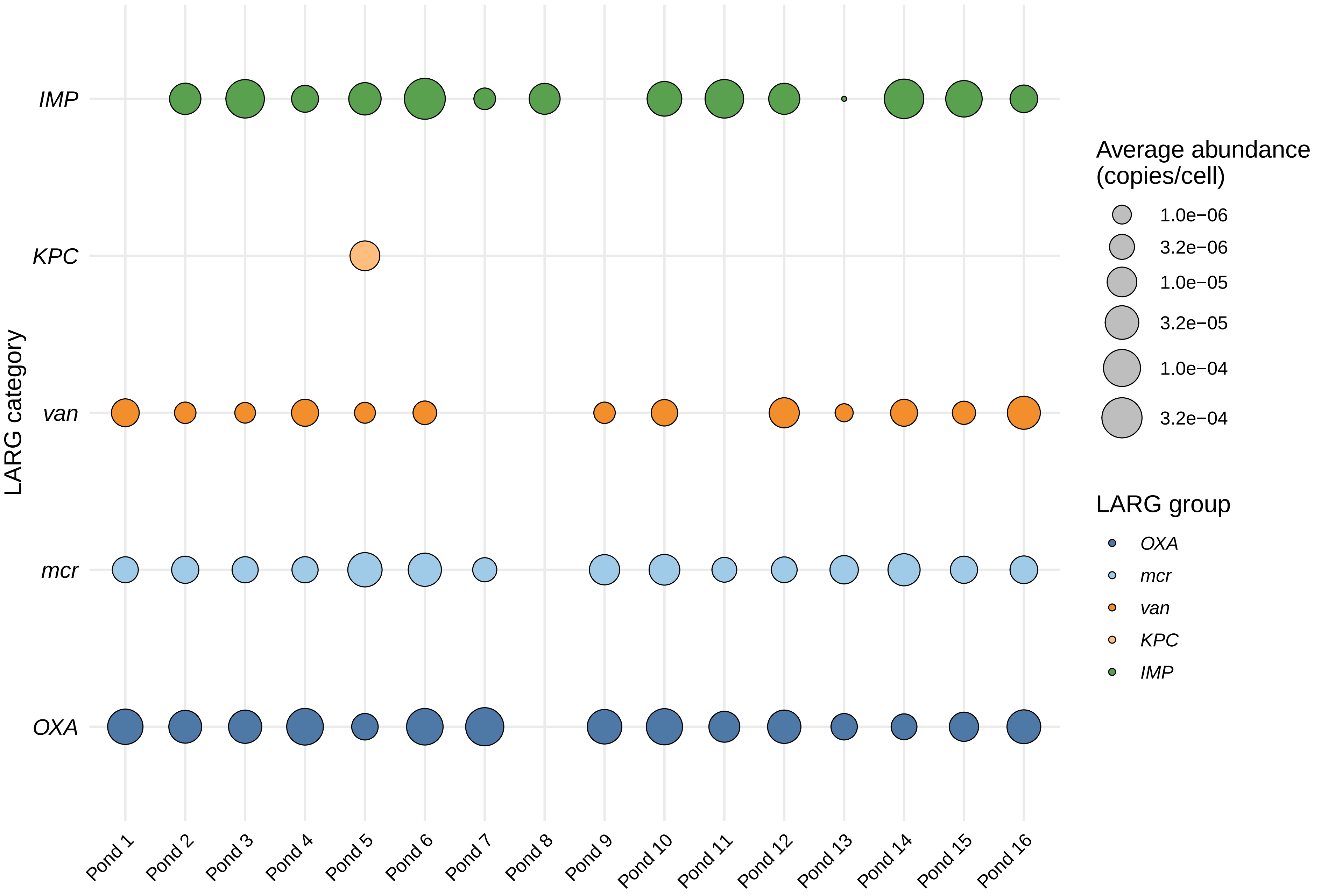

### Supplementary Figure 4

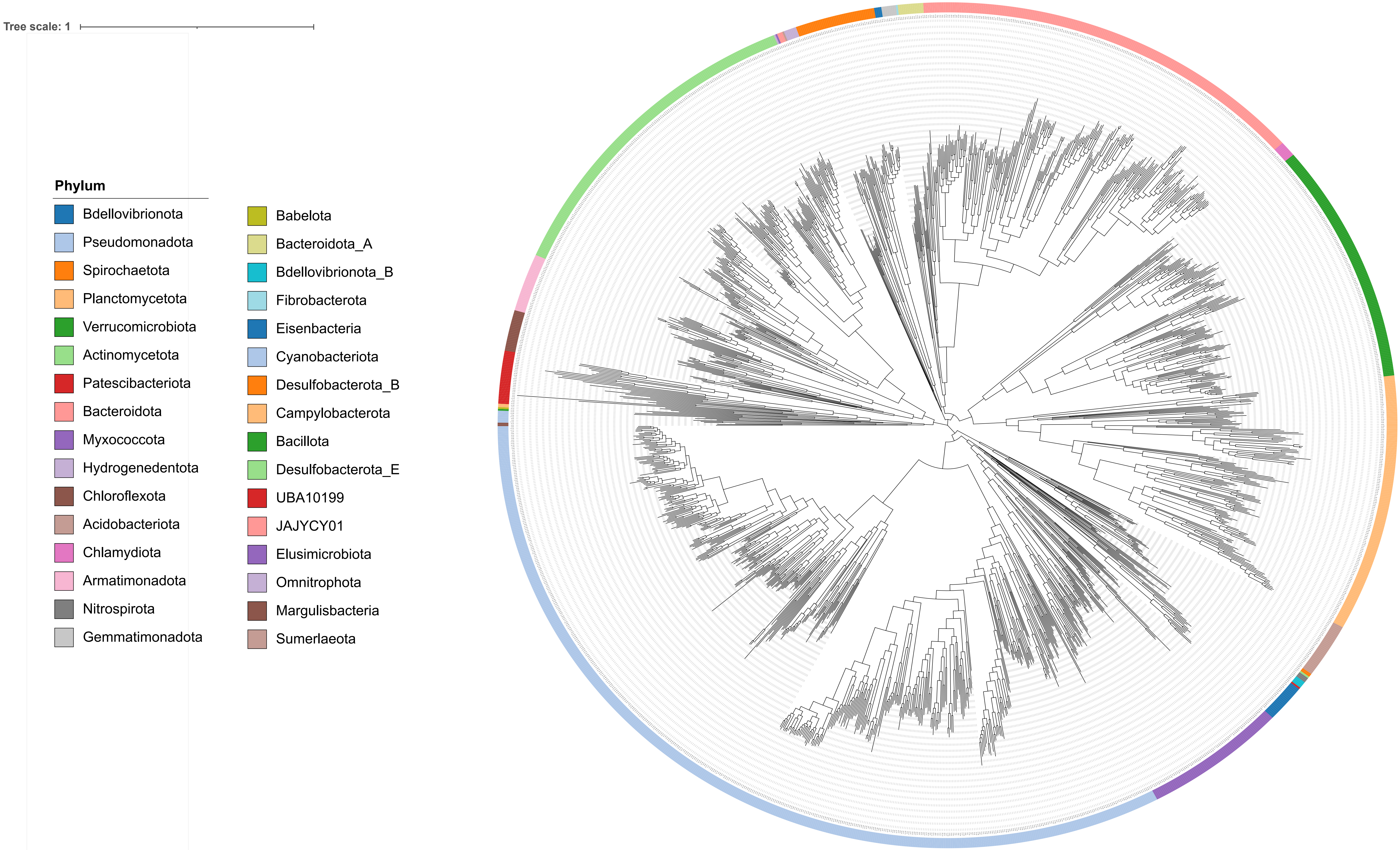

### Supplementary Figure 5

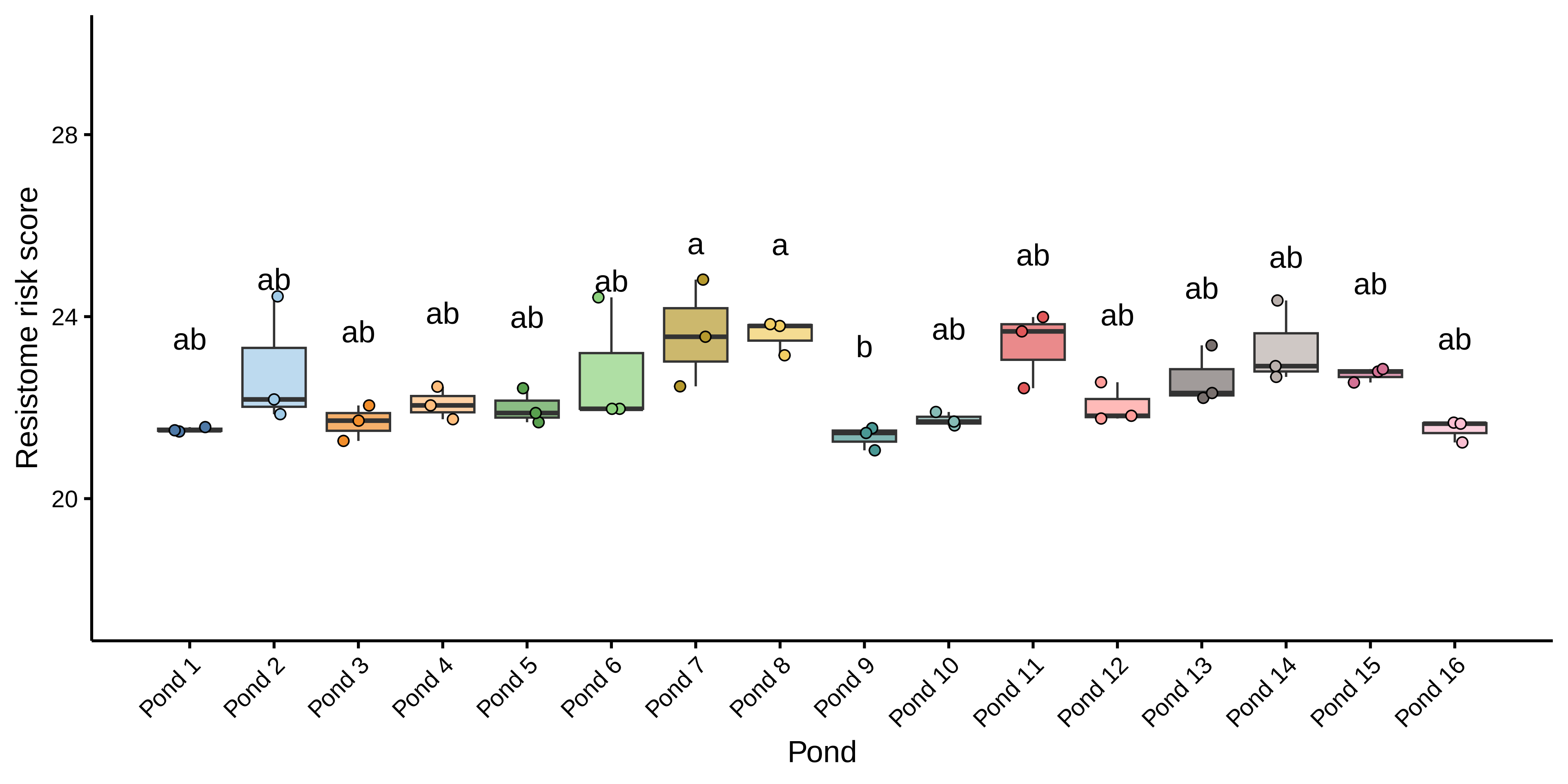

### Supplementary Figure 6

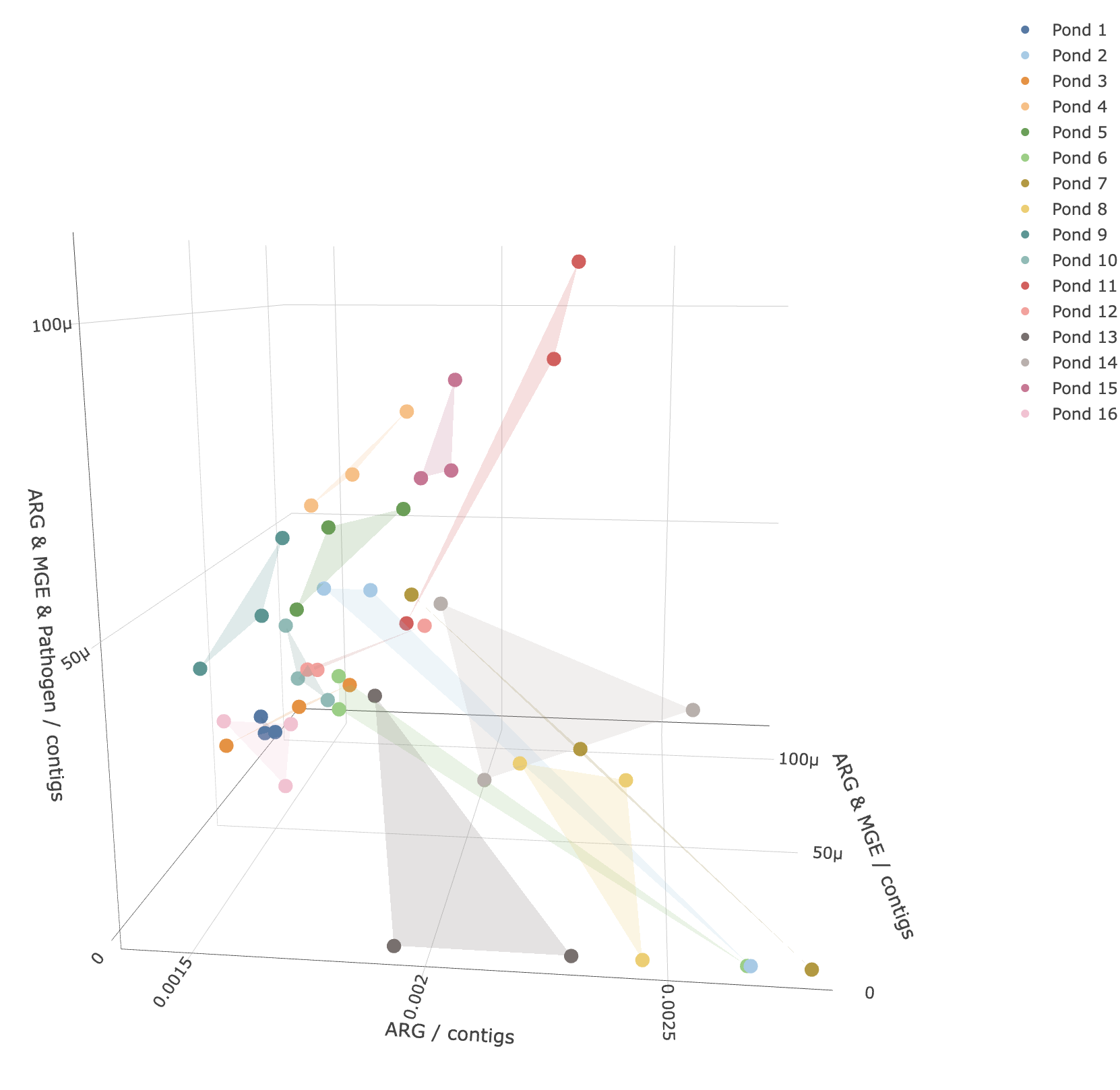
